# Muscle mitochondrial bioenergetic capacities is associated with multimorbidity burden in older adults: the Study of Muscle, Mobility and Aging (SOMMA)

**DOI:** 10.1101/2023.11.06.23298175

**Authors:** Theresa Mau, Terri L. Blackwell, Peggy M. Cawthon, Anthony J. A. Molina, Paul M. Coen, Giovanna Distefano, Philip A. Kramer, Sofhia V. Ramos, Daniel E. Forman, Bret H. Goodpaster, Frederico G. S. Toledo, Kate A. Duchowny, Lauren M. Sparks, Anne B. Newman, Stephen B. Kritchevsky, Steven R. Cummings

## Abstract

**Background:** The geroscience hypothesis posits that aging biological processes contribute to many age-related deficits, including the accumulation of multiple chronic diseases. Though only one facet of mitochondrial function, declines in muscle mitochondrial bioenergetic capacities may contribute to this increased susceptibility to multimorbidity.

**Methods:** The Study of Muscle, Mobility and Aging (SOMMA) assessed *ex vivo* muscle mitochondrial energetics in 764 older adults (mean age =76.4, 56.5% women, 85.9% non-Hispanic white) by high-resolution respirometry of permeabilized muscle fibers. We estimated the proportional odds ratio (POR [95%CI]) for the likelihood of greater multimorbidity (four levels: 0 conditions, N=332; 1 condition, N=299; 2 conditions, N=98; or 3+ conditions, N=35) from an index of 11 conditions, per SD decrement in muscle mitochondrial energetic parameters. Distribution of conditions allowed for testing the associations of maximal muscle energetics with some individual conditions.

**Results:** Lower oxidative phosphorylation supported by fatty acids and/or complex-I and -II linked carbohydrates (e.g., Max OXPHOS_CI+CII_) was associated with a greater multimorbidity index score (POR=1.32[1.13,1.54]) and separately with diabetes mellitus (OR=1.62[1.26,2.09]), depressive symptoms (OR=1.45[1.04,2.00]) and possibly chronic kidney disease (OR=1.57[0.98,2.52]) but not significantly with other conditions (e.g., cardiac arrhythmia, chronic obstructive pulmonary disease).

**Conclusions:** Lower muscle mitochondrial bioenergetic capacities was associated with a worse composite multimorbidity index score. Our results suggest that decrements in muscle mitochondrial energetics may contribute to a greater global burden of disease and is more strongly related to some conditions than others. **(Words** = 233)

## Introduction

Older adults living with more than two chronic conditions—multimorbidity—have higher mortality risk, higher medical costs, and lower healthspan.^5^ Among adults age 65 and older in the United States who are dual eligible for Medicare and Medicaid, the prevalence of multimorbidity was 76.9% in the year 2018.^5^ Geroscience research has focused on identifying discrete biological drivers that underlie multiple chronic aging conditions.^1–4^ It has been posited that mitochondrial dysfunction is a potential biological driver to multimorbidity.^1,6^ Among the multifaceted roles of the mitochondria, mounting evidence has shown that age-related declines in mitochondrial bioenergetic capacities of energetically demanding tissues,^7,8^ such as the skeletal muscle,^9–11^ can greatly impact physical aging health and function.^12–14^

In studies of older adults, *ex vivo* analysis of muscle mitochondrial energetics has been assessed by high resolution respirometry of permeabilized muscle fibers to measure electron transport system (ETS) activity in the presence of complex I- and II-linked carbohydrates and fatty acids. Skeletal muscle mitochondrial energetics have been linked to a range of aging health indicators, including lower-extremity physical function,^13^ mobility,^12,15^ decreased fitness,^13–15^ and frailty.^16,17^ Beyond muscle, a recent study of monocytes from older adults (55-94 years) showed that age-related downregulation of mitochondrial genes associated with multimorbidity burden.^18^ Age-associated decreases in mitochondrial bioenergetic capacities have been reported in multiple tissues: these organism wide decrements support the notion that it could majorly contribute to age-related diseases.^19^

Abnormalities of skeletal muscle mitochondrial bioenergetics in patients with chronic heart failure,^20–23^ type 2 diabetes,^24,25^ and chronic kidney disease^26,27^ have been reported. In some of these studies,^20,22,26^ among participants with the same condition, those with lower muscle energetics tended to exhibit greater disease severity and physical dysfunction than those without a chronic condition. Reduced physical activity and elevated adiposity in older age are also associated with lower mitochondrial energetics and cardiorespiratory fitness.^28–30^ Recent studies,^13,31^ including ours in the Study of Muscle, Mobility and Aging (SOMMA),^32^ demonstrated that independent of physical activity and adiposity, lower maximal muscle mitochondrial energetics was associated with poorer cardiorespiratory fitness and physical function. In general, the current literature describes association between mitochondrial energetics and individual diseases, conditions, or decrements. However, the direct relation of muscle energetics with multimorbidity within the context of a well-characterized cohort with gold-standard measures of skeletal muscle mitochondrial bioenergetics has not been investigated.

The primary aim of this report is to assess whether skeletal muscle mitochondrial energetics are associated with multimorbidity burden in older adults participating in SOMMA. The age-related conditions selected in our multimorbidity index occur in energetically demanding tissues (e.g., skeletal muscle, kidney, heart);^19^ therefore, we hypothesized that older adults with lower muscle mitochondrial energetics would have greater multimorbidity burden. We also investigated the association of skeletal muscle mitochondrial energetics with individual conditions in the multimorbidity index, where the prevalence of the condition was sufficiently high to allow for power to detect associations.

## Online Methods

### Study Cohort

The Study of Muscle, Mobility and Aging (SOMMA) is a prospective longitudinal cohort study to determine the biological basis of human aging with emphasis on mobility decline. Recruitment and baseline assessments (i.e., enrollment criteria, clinical measures, primary outcomes) between April 2019 and December 2021 of 879 adults aged 70 or older at the University of Pittsburgh and Wake Forest University School of Medicine have been previously described.^32^ Briefly, individuals were eligible to participate if they were ≥70 years old, willing and able to complete a skeletal muscle biopsy and undergo magnetic resonance (MR). Participants must have been able to complete the 400-meter (400m) walk. Individuals who appeared as they might not be able to complete the 400m walk at the in-person screening visit completed a short distance walk (4 meters) to ensure their walking speed was ≥0.6 m/s. They were excluded if they had body mass index (BMI) ≥40 kg/m^2^; had an active malignancy or dementia; or any medical contraindication to biopsy or MR. All participants provided written informed consent. SOMMA was approved by the Western IRB-Copernicus Group (WCG) Institutional Review Board (#20180764).

### Baseline general characteristic measures

At baseline, participants completed questionnaires and exams over three clinic visits. The three visits were conducted within six to eight weeks from the first visit. SOMMA baseline questionnaires collected a range of self-reported measures, including date of birth, sex, race (Non-Hispanic White or racial/ethnic minority individual), education (i.e., <high school, some college, college graduate, or post college work), alcohol intake (drinks/week), smoking history (no, past, current), and medical history. Participants provided an inventory of prescription containers, and we totaled medications taken in the last 30 days. The 10-item version of the Center of Epidemiologic Studies Depression Scale (CESD-10) assessed depressive symptoms (score ≥10, range=0-30).^57^ Height (m) was measured on stadiometers and weight (kg) on digital scales, and body mass index (BMI) was calculated (kg/m^2^). Whole-body MR scans and Dixon water-fat imaging were processed using AMRA Researcher (AMRA Medical AB, Linköping, Sweden), and we calculated total abdominal subcutaneous adipose tissue (ASAT) and visceral adipose tissue (VAT) volumes (L).^58^ Whole body skeletal muscle mass (kg) was assessed by deuterated creatine (D_3_Cr) dilution method and corrected for spillage.^59,60^ Objectively-measured physical activity was assessed with a wrist-worn Actigraph GT9X for 7-full days. We calculated the total time (min/day) spent in moderate-to-vigorous physical activity (MVPA), averaged over all days they wore the device.^62,63^

### Baseline physical function measures

Participant eligibility required the ability to complete a 400-meter (400m) walk within 15 minutes. The 400m walk was assessed in a 40-meter course at the participant’s usual or preferred pace for 10 laps without assistive devices except for a straight cane.^64,65^ Total 400m walk speed was calculated as an average (meters/second) which included rest time if participant chose to pause during the test. The isometric hand grip strength from both hands were assessed in duplicate with an adjustable hydraulic dynamometer (Jamar); the maximal value across both hands was analyzed.^66,67^ Participants completed a standardized cardiopulmonary exercise test (CPET) using a modified Balke or manual protocol to measure ventilatory gases during exercise to estimate VO_2peak_ (mL/kg/min) as previously described.^68^

### SOMMA multimorbidity index scores

Many definitions of multimorbidity exist. To construct the multimorbidity index in SOMMA, we curated a list of eleven age-related chronic conditions that closely follows the Rochester Epidemiology Project (REP).^69^ The REP is a population-based records-linkage system in Rochester, Minnesota that has been used extensively to determine the prevalence and incidence of multimorbidity across different ages, sex, and ethnicities.^69^ Based on self-reported diagnosis of common medical conditions, we calculated a SOMMA Multimorbidity Score using a modified list of chronic conditions from the REP. The score included eleven age-related conditions: cancer, chronic kidney disease or renal failure, atrial fibrillation, lung disease (i.e., chronic obstructive pulmonary disease, bronchitis, asthma, or emphysema), coronary heart disease (i.e., blocked artery or myocardial infarction), depressive symptoms (e.g., ≥10 score on CESD-10 consistent with depressive symptoms), heart failure, dementia, diabetes, stroke, and aortic stenosis. The REP included arthritis and osteoporosis which we omitted, as diagnoses of these highly reported conditions are not readily confirmed after physician preliminary diagnosis. Non-melanoma skin cancer was excluded from the cancer group. Participants were grouped into the number of chronic conditions they reported at baseline visit, which was either zero, one, two, and three or more conditions.

### High resolution respirometry of permeabilized muscle fibers

Under local anesthesia, a percutaneous biopsy of the middle region of the vastus lateralis was collected using a Bergstrom canula with suction;^70^ permeabilized muscle fiber bundles were prepared as previously described and underwent high-resolution respirometry. We used two standardized protocols,^13,15^ run in duplicate, to assess mitochondrial respiratory capacity.

Permeabilized muscle fiber bundles were transferred into an Oroboros Oxygraph-2k (O2k, Oroboros Instruments, Innsbruck, Austria). Respiration chambers were maintained at 37°C, and O_2_ concentration kept between 400-200µM. In protocol 1, complex I-supported leak respiration (Leak_CI_ or State 4) was determined with the addition of pyruvate (5mM) and malate (2mM).

Sequential additions of adenosine diphosphate (ADP) induced complex I–supported oxidative phosphorylation (Submax OXPHOS_CI_ (225uM ADP) and OXPHOS_CI_ (4.2mM ADP)). Glutamate (10mM) was added to determine maximal complex I-supported OXPHOS (Max OXPHOS_CI_) and succinate (10mM) to measure maximal complex I- and II-supported OXPHOS (Max OXPHOS_CI+CII_ or State 3). Carbonyl cyanide-p-trifluoromethoxyphenylhydrazone (FCCP) was titrated (0.5-3.0µM) to determine maximal complex I- and II-supported electron transport system capacity or uncoupled respiration (Max ETS_CI+CII_) capacity. In protocol 2, FA (fatty acid)-supported leak respiration (Leak_CI+FAO_) was determined through the addition of palmitoylcarnitine (25μM) and malate (2mM). ADP (4.0mM) was added to elicit complex I and FAO (Fatty acid oxidation)-supported OXPHOS (OXPHOS_CI+FAO_). Glutamate (10mM) was then added to determine maximal complex I and FAO-supported maximal OXPHOS (Max OXPHOS_CI+FAO_). Finally, succinate (10mM) was added to stimulate maximal CI+II and FAO-supported OXPHOS (Max OXPHOS_CI+CII+FAO_). Cytochrome c (10uM) was added to both protocols to assess the integrity of the outer mitochondrial membrane, and any sample with a response higher than 15% were not included in the results. All respirometry data were analyzed with Datlab 7.4 and normalized to the wet weight of permeabilized muscle fibers. Respiratory control ratio (RCR), defined as the ratio of ADP stimulated respiration to non-ADP respiration, was 7.1 ± 2.9 (mean ± SD) for protocol 1 (N=730) and 3.5 ± 1.7 for protocol 2 (N=538). A summary of all respirometry variables is shown in **Supplementary Table 1**.

### Statistical Analyses

Characteristics and muscle mitochondrial energetic parameters were summarized across the 4-category SOMMA Multimorbidity Index (0 conditions, 1 condition, 2 conditions, and 3 or more conditions). For testing the linear trend of characteristics across multimorbidity score categories, we used linear regression for normally distributed continuous variables and two-sided Jonckheere-Terpstra for skewed or categorical variables. We tested for sex interactions (all *p-interaction* ≥0.17) with all muscle mitochondrial energetics (respirometry) variables by modeling the main effects (e.g., sex and Max OXPHOS_CI+CII_) and their product (e.g., sex*Max OXPHOS_CI+CII_) together.

We estimated the proportional odds ratio (POR) for greater multimorbidity scores expressed as POR per 1 standard deviation (SD) decrement of muscle mitochondrial energetics. The proportionality assumption was met for all models which allowed us to report a singular OR that reflects the likelihood of being in a greater multimorbidity category. Each measure of muscle mitochondrial energetics was assessed in a separate model. The associations of the individual components of the multimorbidity index were outcomes in separate logistic models, shown per one SD decrement of muscle mitochondrial energetics. For all associations examined, two sets of adjusted models were reported. The minimally adjusted model adjusted for technician, age, and sex. The fully adjusted model adjusted for technician, age, sex, race, education, smoking, alcohol, weight*height, adiposity (ASAT, VAT), and objectively measured moderate-to-vigorous physical activity. For associations of individual multimorbidity index conditions with maximal muscle mitochondrial energetics, we only reported the odds ratio (95% CI) from a minimally adjusted logistic model (age, sex, and site) to ensure sample size and power would not decrease from missing covariates. All significance levels reported were 2-sided, and all statistical analyses were performed using SAS software, version 9.4 (SAS Institute, Inc, Cary, NC). Some figures were generated in R version 4.01.

### Analysis sample

Of the 879 participants with complete baseline measures (**Supplementary Figure 1**), only a few were missing data on one or more self-reported conditions (N=5) or refused the CES-D (N=9). In total, 764 participants had data for both the SOMMA Multimorbidity Index scores and at least 1 measure of muscle mitochondrial energetics either protocol 1 or 2; with 733 participants having Protocol 1 data, and 540 participants having Protocol 2 data.

## Results

The majority (85.9%) of SOMMA participants were non-Hispanic White, and the average age of these participants (56.5% women) was 76.4 years (**Table 1**). Participants were grouped into four categories of multimorbidity scores. At baseline, 43.5% of participants had 0 conditions, 39.1% had 1 condition, 12.8% had 2 conditions, and 4.6% had 3 or more of 11 conditions (**Table 2**).

**Table 1.**
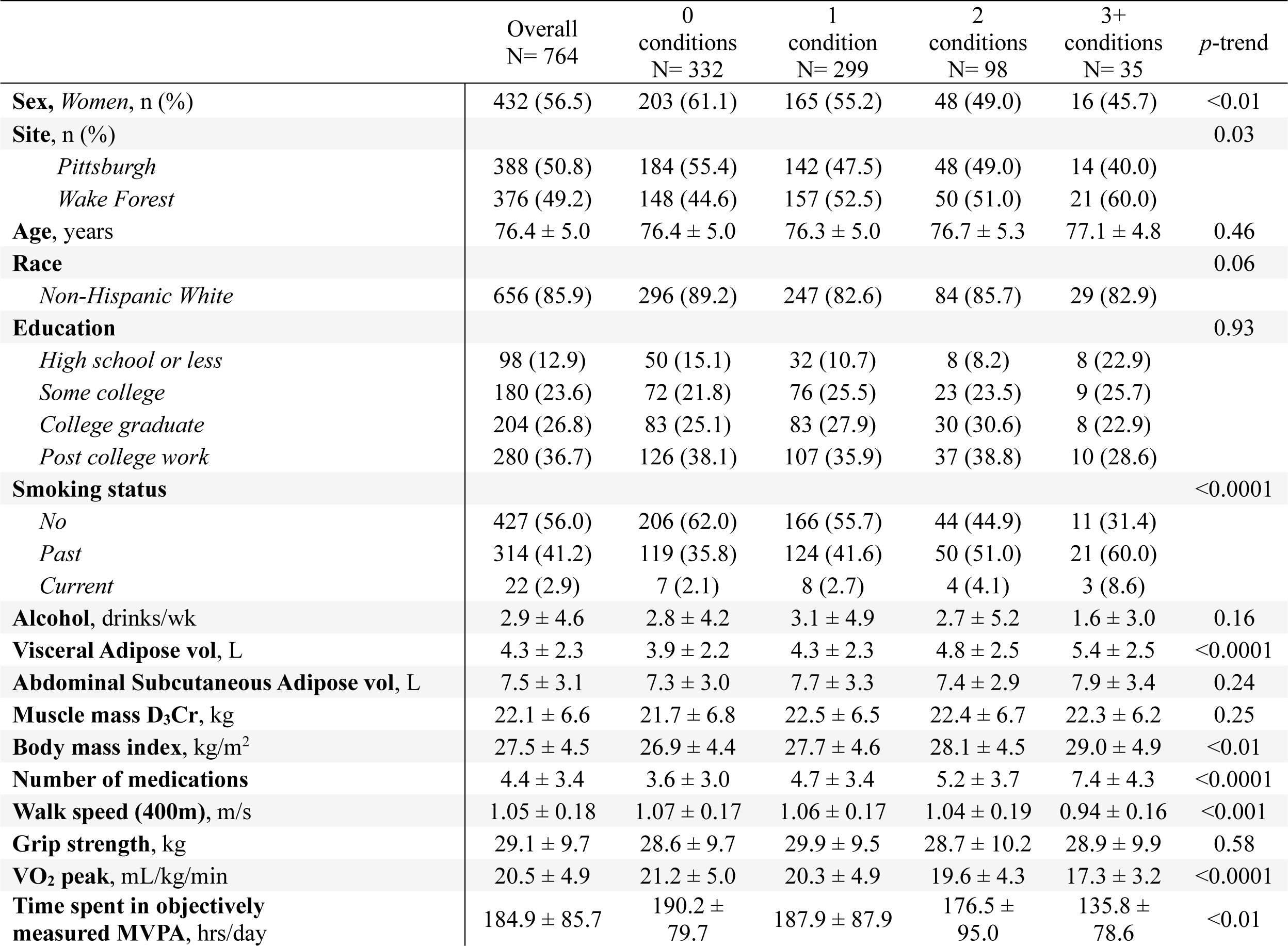
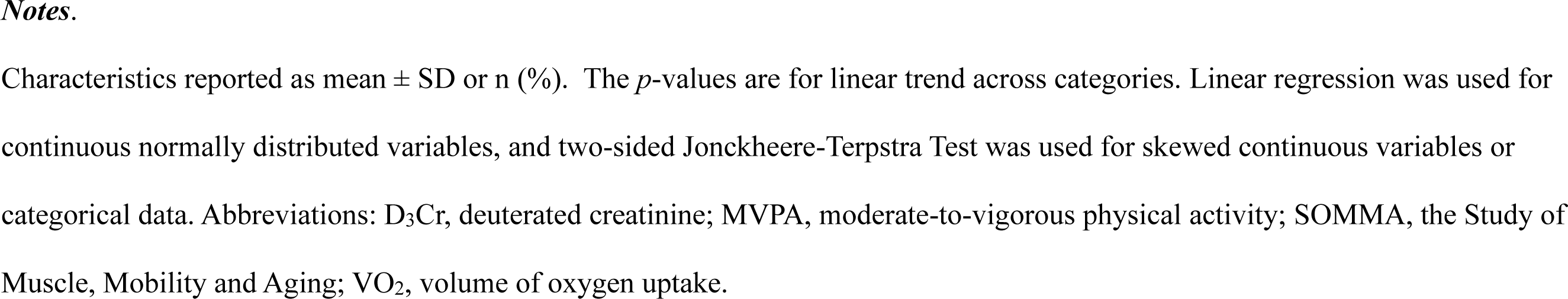
Baseline characteristics of SOMMA older adults (age≥70 years) by multimorbidity index scores.

|  | Overall<br>N= 764 | 0<br>conditions<br>N= 332 | 1<br>condition<br>N= 299 | 2<br>conditions<br>N= 98 | 3+<br>conditions<br>N= 35 | *p*-trend |
| --- | --- | --- | --- | --- | --- | --- |
| **Sex, Women, n (%)** | 432 (56.5) | 203 (61.1) | 165 (55.2) | 48 (49.0) | 16 (45.7) | <0.01 |
| **Site, n (%)** |  |  |  |  |  | 0.03 |
| *Pittsburgh* | 388 (50.8) | 184 (55.4) | 142 (47.5) | 48 (49.0) | 14 (40.0) |  |
| *Wake Forest* | 376 (49.2) | 148 (44.6) | 157 (52.5) | 50 (51.0) | 21 (60.0) |  |
| **Age, years** | 76.4 ± 5.0 | 76.4 ± 5.0 | 76.3 ± 5.0 | 76.7 ± 5.3 | 77.1 ± 4.8 | 0.46 |
| **Race** |  |  |  |  |  | 0.06 |
| *Non-Hispanic White* | 656 (85.9) | 296 (89.2) | 247 (82.6) | 84 (85.7) | 29 (82.9) |  |
| **Education** |  |  |  |  |  | 0.93 |
| *High school or less* | 98 (12.9) | 50 (15.1) | 32 (10.7) | 8 (8.2) | 8 (22.9) |  |
| *Some college* | 180 (23.6) | 72 (21.8) | 76 (25.5) | 23 (23.5) | 9 (25.7) |  |
| *College graduate* | 204 (26.8) | 83 (25.1) | 83 (27.9) | 30 (30.6) | 8 (22.9) |  |
| *Post college work* | 280 (36.7) | 126 (38.1) | 107 (35.9) | 37 (38.8) | 10 (28.6) |  |
| **Smoking status** |  |  |  |  |  | <0.0001 |
| *No* | 427 (56.0) | 206 (62.0) | 166 (55.7) | 44 (44.9) | 11 (31.4) |  |
| *Past* | 314 (41.2) | 119 (35.8) | 124 (41.6) | 50 (51.0) | 21 (60.0) |  |
| *Current* | 22 (2.9) | 7 (2.1) | 8 (2.7) | 4 (4.1) | 3 (8.6) |  |
| **Alcohol, drinks/wk** | 2.9 ± 4.6 | 2.8 ± 4.2 | 3.1 ± 4.9 | 2.7 ± 5.2 | 1.6 ± 3.0 | 0.16 |
| **Visceral Adipose vol, L** | 4.3 ± 2.3 | 3.9 ± 2.2 | 4.3 ± 2.3 | 4.8 ± 2.5 | 5.4 ± 2.5 | <0.0001 |
| **Abdominal Subcutaneous Adipose vol, L** | 7.5 ± 3.1 | 7.3 ± 3.0 | 7.7 ± 3.3 | 7.4 ± 2.9 | 7.9 ± 3.4 | 0.24 |
| **Muscle mass D<sub>3</sub>Cr, kg** | 22.1 ± 6.6 | 21.7 ± 6.8 | 22.5 ± 6.5 | 22.4 ± 6.7 | 22.3 ± 6.2 | 0.25 |
| **Body mass index, kg/m<sup>2</sup>** | 27.5 ± 4.5 | 26.9 ± 4.4 | 27.7 ± 4.6 | 28.1 ± 4.5 | 29.0 ± 4.9 | <0.01 |
| **Number of medications** | 4.4 ± 3.4 | 3.6 ± 3.0 | 4.7 ± 3.4 | 5.2 ± 3.7 | 7.4 ± 4.3 | <0.0001 |
| **Walk speed (400m), m/s** | 1.05 ± 0.18 | 1.07 ± 0.17 | 1.06 ± 0.17 | 1.04 ± 0.19 | 0.94 ± 0.16 | <0.001 |
| **Grip strength, kg** | 29.1 ± 9.7 | 28.6 ± 9.7 | 29.9 ± 9.5 | 28.7 ± 10.2 | 28.9 ± 9.9 | 0.58 |
| **VO<sub>2</sub> peak, mL/kg/min** | 20.5 ± 4.9 | 21.2 ± 5.0 | 20.3 ± 4.9 | 19.6 ± 4.3 | 17.3 ± 3.2 | <0.0001 |
| **Time spent in objectively measured MVPA, hrs/day** | 184.9 ± 85.7 | 190.2 ± 79.7 | 187.9 ± 87.9 | 176.5 ± 95.0 | 135.8 ± 78.6 | <0.01 |

Characteristics reported as mean ± SD or n (%). The *p*-values are for linear trend across categories. Linear regression was used for continuous normally distributed variables, and two-sided Jonckheere-Terpstra Test was used for skewed continuous variables or categorical data. Abbreviations: D<sub>3</sub>Cr, deuterated creatinine; MVPA, moderate-to-vigorous physical activity; SOMMA, the Study of Muscle, Mobility and Aging; VO<sub>2</sub>, volume of oxygen uptake.

**Table 2.**
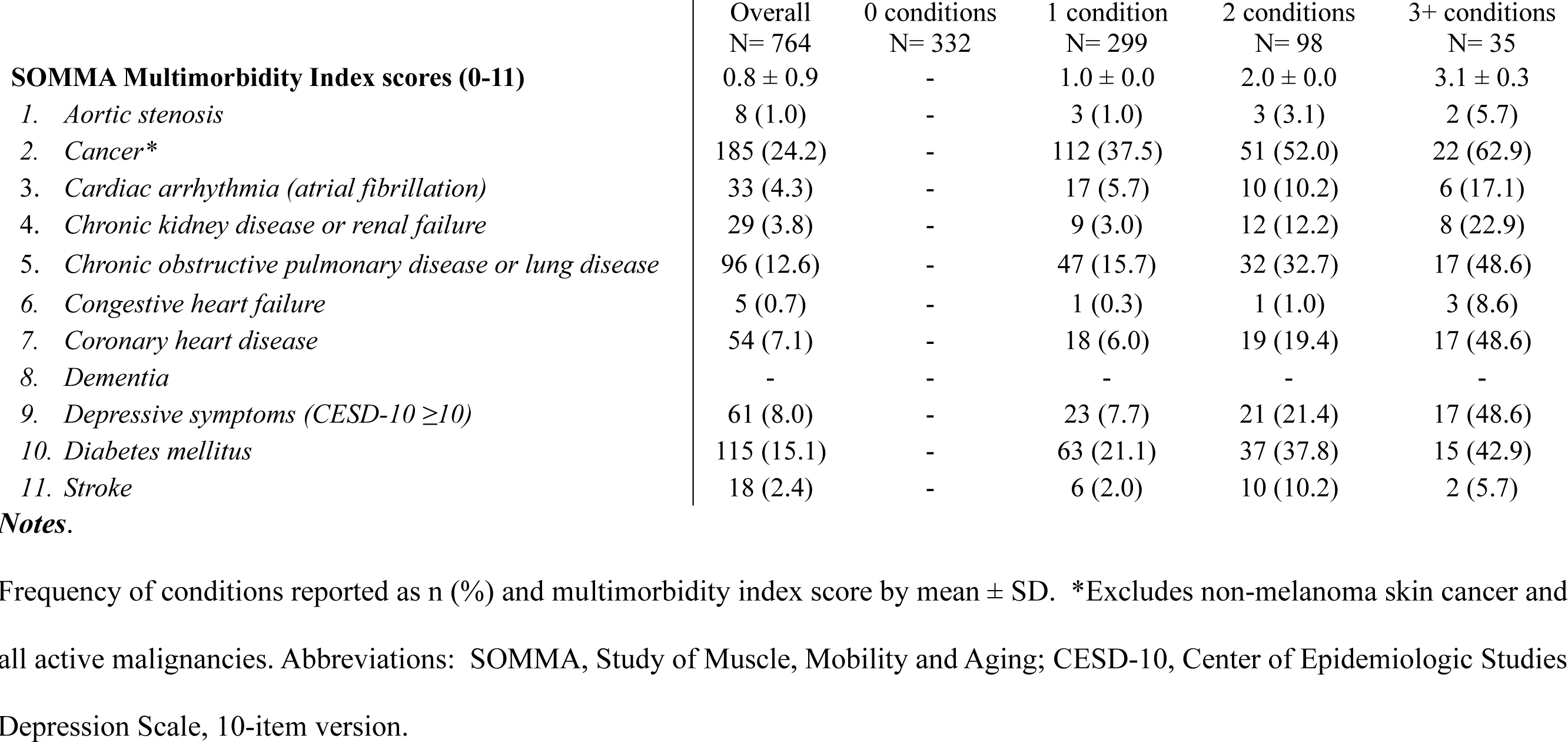
Frequency of chronic medical conditions by total number of conditions SOMMA participants reported.

|  | Overall<br>N= 764 | 0 conditions<br>N= 332 | 1 condition<br>N= 299 | 2 conditions<br>N= 98 | 3+ conditions<br>N= 35 |
| --- | --- | --- | --- | --- | --- |
| **SOMMA Multimorbidity Index scores (0-11)** | 0.8 ± 0.9 | - | 1.0 ± 0.0 | 2.0 ± 0.0 | 3.1 ± 0.3 |
| 1. *Aortic stenosis* | 8 (1.0) | - | 3 (1.0) | 3 (3.1) | 2 (5.7) |
| 2. *Cancer\** | 185 (24.2) | - | 112 (37.5) | 51 (52.0) | 22 (62.9) |
| 3. *Cardiac arrhythmia (atrial fibrillation)* | 33 (4.3) | - | 17 (5.7) | 10 (10.2) | 6 (17.1) |
| 4. *Chronic kidney disease or renal failure* | 29 (3.8) | - | 9 (3.0) | 12 (12.2) | 8 (22.9) |
| 5. *Chronic obstructive pulmonary disease or lung disease* | 96 (12.6) | - | 47 (15.7) | 32 (32.7) | 17 (48.6) |
| 6. *Congestive heart failure* | 5 (0.7) | - | 1 (0.3) | 1 (1.0) | 3 (8.6) |
| 7. *Coronary heart disease* | 54 (7.1) | - | 18 (6.0) | 19 (19.4) | 17 (48.6) |
| 8. *Dementia* | - | - | - | - | - |
| 9. *Depressive symptoms (CESD-10 ≥10)* | 61 (8.0) | - | 23 (7.7) | 21 (21.4) | 17 (48.6) |
| 10. *Diabetes mellitus* | 115 (15.1) | - | 63 (21.1) | 37 (37.8) | 15 (42.9) |
| 11. *Stroke* | 18 (2.4) | - | 6 (2.0) | 10 (10.2) | 2 (5.7) |

**Notes.**
Frequency of conditions reported as n (%) and multimorbidity index score by mean ± SD. \*Excludes non-melanoma skin cancer and all active malignancies. Abbreviations: SOMMA, Study of Muscle, Mobility and Aging; CESD-10, Center of Epidemiologic Studies Depression Scale, 10-item version.

Participants with a greater number of chronic conditions were generally not older in age but had slower gait speed and lower cardiorespiratory fitness (VO_2peak_). They engaged in less time spent in moderate-to-vigorous physical activity (MVPA) and had higher body mass index (BMI), visceral adiposity, and used more prescription medications. Muscle mass and grip strength did not differ among the multimorbidity groups. We found no sex interactions (all interactions *p*≥0.17) for muscle mitochondrial respirometry parameters and chronic conditions in all the statistical models we implemented. Of the eleven age-associated chronic conditions in the multimorbidity index, the most common was (**Figure 1**): cancer (24.2%), followed by diabetes mellitus (15.1%), chronic obstructive pulmonary disease or lung disease (12.6%), depressive symptoms (8.0%), coronary heart disease (7.1%), cardiac arrhythmia or atrial fibrillation (4.3%), chronic kidney disease or renal failure (3.8%), stroke (2.4%), aortic stenosis (1.0%), congestive heart failure (0.7%), and dementia (excluded at enrollment; 0%).

**Figure 1.**
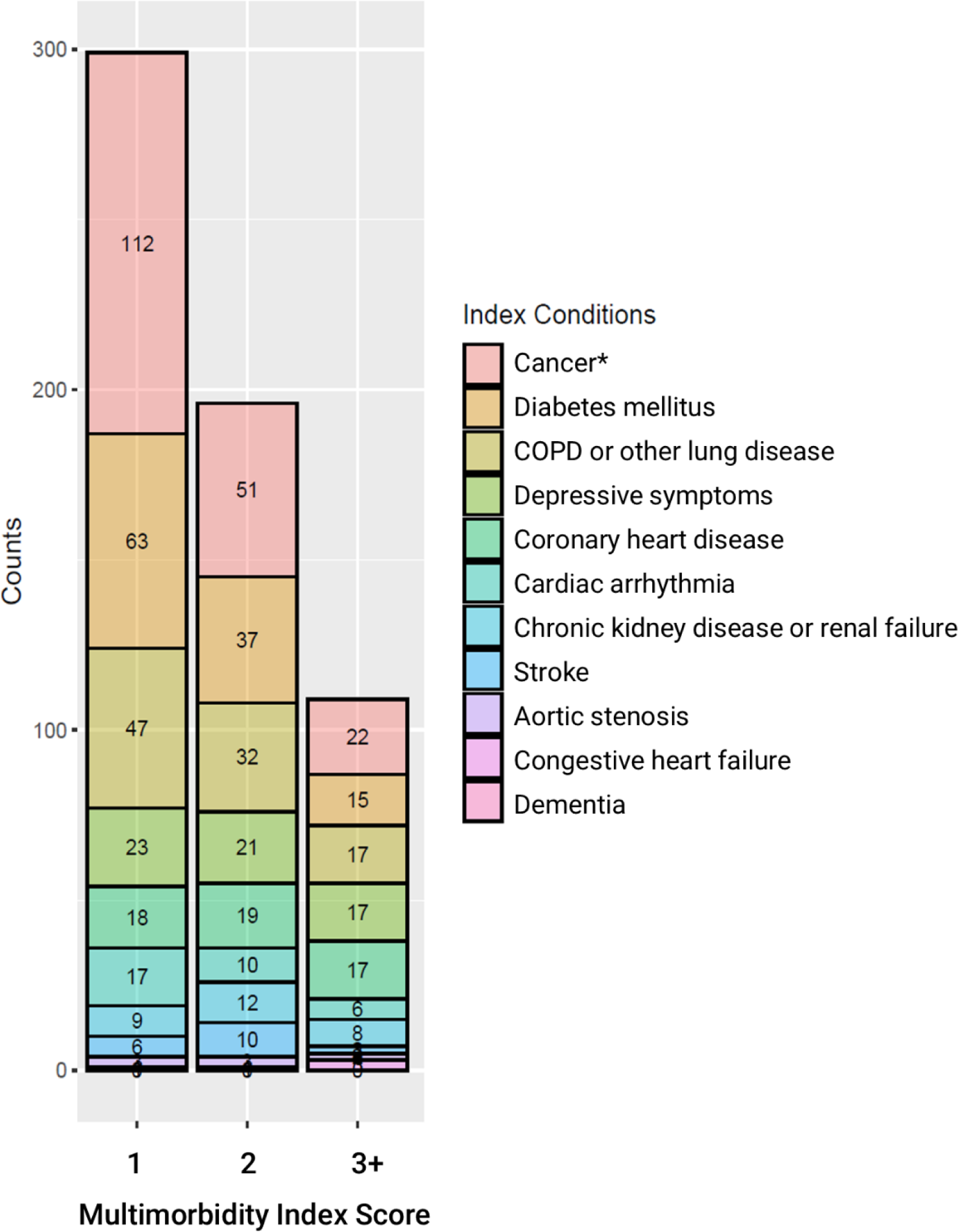
Distribution of age-related conditions in the SOMMA Multimorbidity Index. The count of each chronic medical condition is plotted by the total number of conditions participants reported. The legend (right) corresponds to the highest to least frequent conditions in the multimorbidity index. *Cancer excludes non-melanoma skin cancer and all active malignancies.

On average, lower means of muscle mitochondrial energetics were related to greater multimorbidity index scores (**Figure 2**). This was true for all respirometry measures except Submax OXPHOS_CI_ (**Figure 2A**) and Leak_CI+FAO_ (**Figure 2B**). For every 1 SD decrement (POR [95% CI]) in each muscle mitochondrial energetic parameter (**Figure 3**), older adults were more likely to have a greater multimorbidity index score (e.g., Max OXPHOS_CI+CII_, POR=1.32[1.13, 1.54]; Max ETS, POR=1.30[1.11,1.53]). These associations remained significant in fully adjusted models (e.g., Max OXPHOS_CI+CII_, POR=1.25[1.06,1.48]; Max ETS, POR=1.25[1.04,1.49]), except for Submax OXPHOS_CI_ (POR=1.08[0.90,1.28]) (**Figure 3A**), Leak_CI+FAO_ (POR=1.01[0.83,1.23]), Max OXPHOS_CI+FAO_ (POR=1.14[0.94,1.38]), and Max OXPHOS_CI+CII+FAO_ (POR=1.22[0.99,1.50]) (**Figure 3B**).

**Figure 2.**
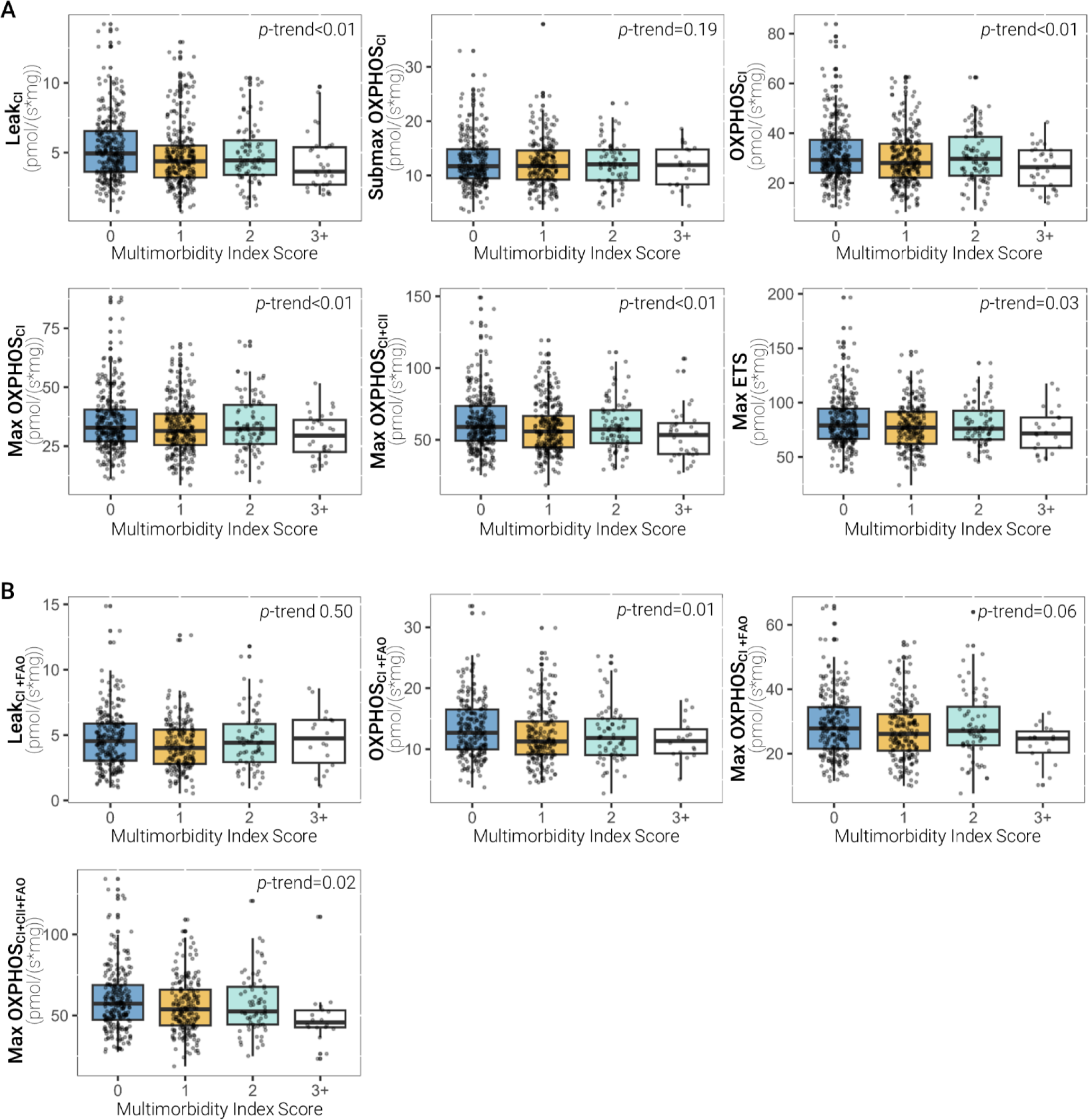
Muscle mitochondrial bioenergetic capacities by multimorbidity index scores in SOMMA older adults. Boxplots of each measure of muscle mitochondrial respiration (pmol/(s*mg)) in the presence of **A**) carbohydrate fuels and **B**) fatty and carbohydrate fuels are shown with the *p*-trend across scores reported.

**Figure 3.**
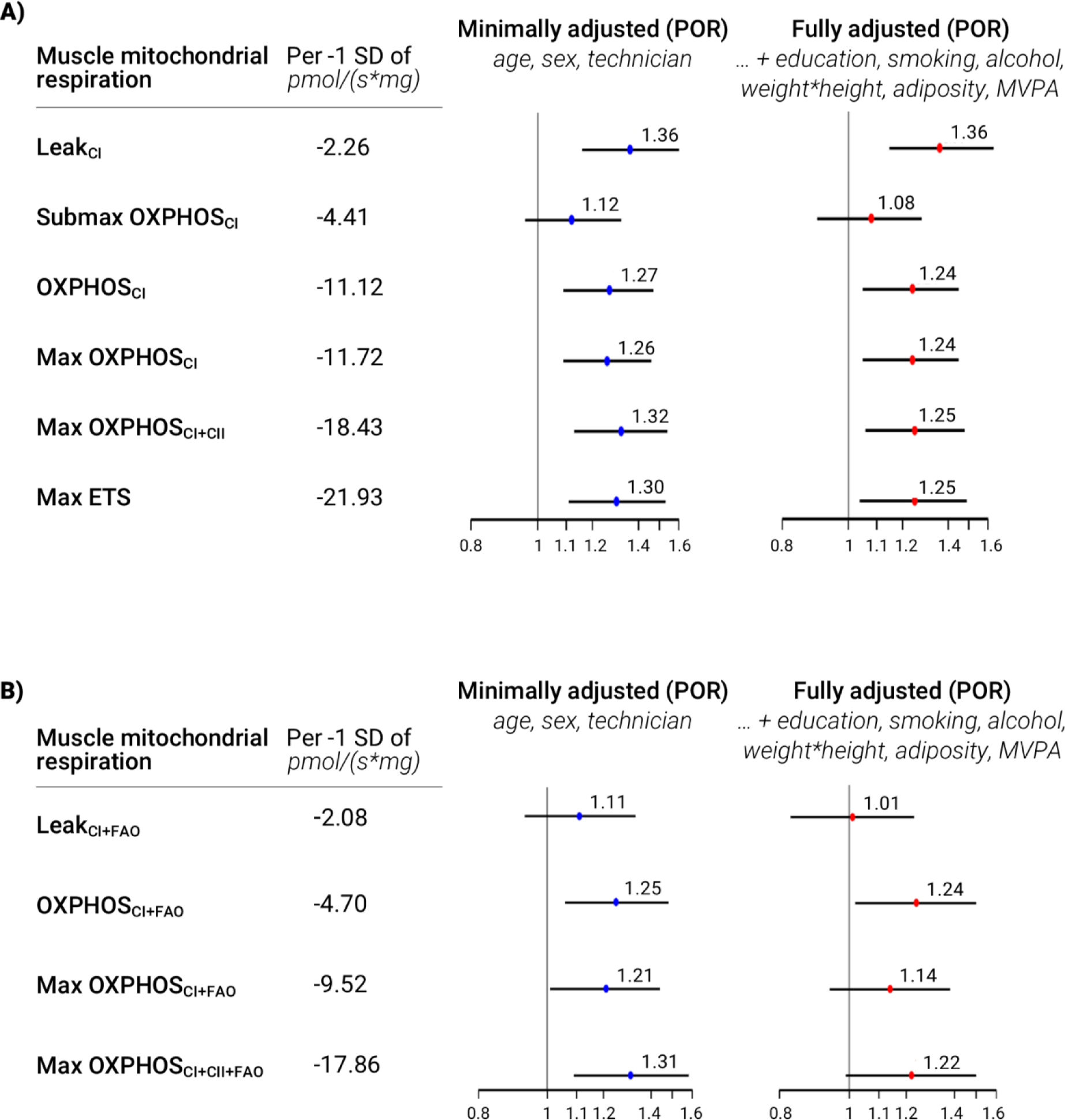
Lower muscle mitochondrial energetics is associated with higher odds of greater multimorbidity burden in SOMMA. Measures of association are proportional odds ratios (POR) where the likelihood of greater frailty is reported per 1 standard deviation (SD) decrement of **A**) carbohydrate-supported and **B**) carbohydrate and fatty-acid-supported muscle mitochondrial respiration. For variable names, refer to **Supplementary Table 1**.

The low prevalence for some of the individual conditions from the multimorbidity index did not allow for assessing the associations of muscle mitochondrial energetics with all conditions individually. SOMMA excluded individuals who reported a dementia diagnosis, so while we included this condition in our index; no participants had dementia at baseline. In addition, only a few participants reported a diagnosis history of congestive heart failure (N=5), aortic stenosis (N=8), and stroke (N=18); this may be due to the exclusion criteria which restricted enrollment of those with severe disease. All other conditions were examined as separate outcomes in minimally adjusted models to assess for associations of maximal muscle mitochondrial energetics with the specific condition (**Figure 4**). We found that those with lower Max OXPHOS_CI+CII_ had higher odds of having depressive symptoms (OR=1.45[1.04,2.00]), diabetes mellitus (OR=1.62[1.26,2.09]), and possibly chronic kidney disease or renal failure (OR=1.57[0.98,2.52]). Similarly, those with lower Max OXPHOS_CI+CII+FAO_ also had higher odds of having depressive symptoms (OR=1.78[1.18, 2.68]) and diabetes mellitus (OR=1.54[1.16, 2.04]) but the association with chronic kidney disease was not significant (OR=1.50[0.86, 2.63]) (**Figure 4**). Max OXPHOS_CI+CII_ had a borderline association with coronary heart disease (OR=1.26[0.92,1.74]), and there was no significant association of Max OXPHOS_CI+CII+FAO_ (OR=0.88[0.60, 1.30]) with coronary heart disease. All other conditions assessed had similar associations, where each SD decrement of Max OXPHOS_CI+CII_ or Max OXPHOS_CI+CII+FAO_ had an increased likelihood of disease (OR>1.0), but the confidence interval ranges were wide and included 1.

**Figure 4.**
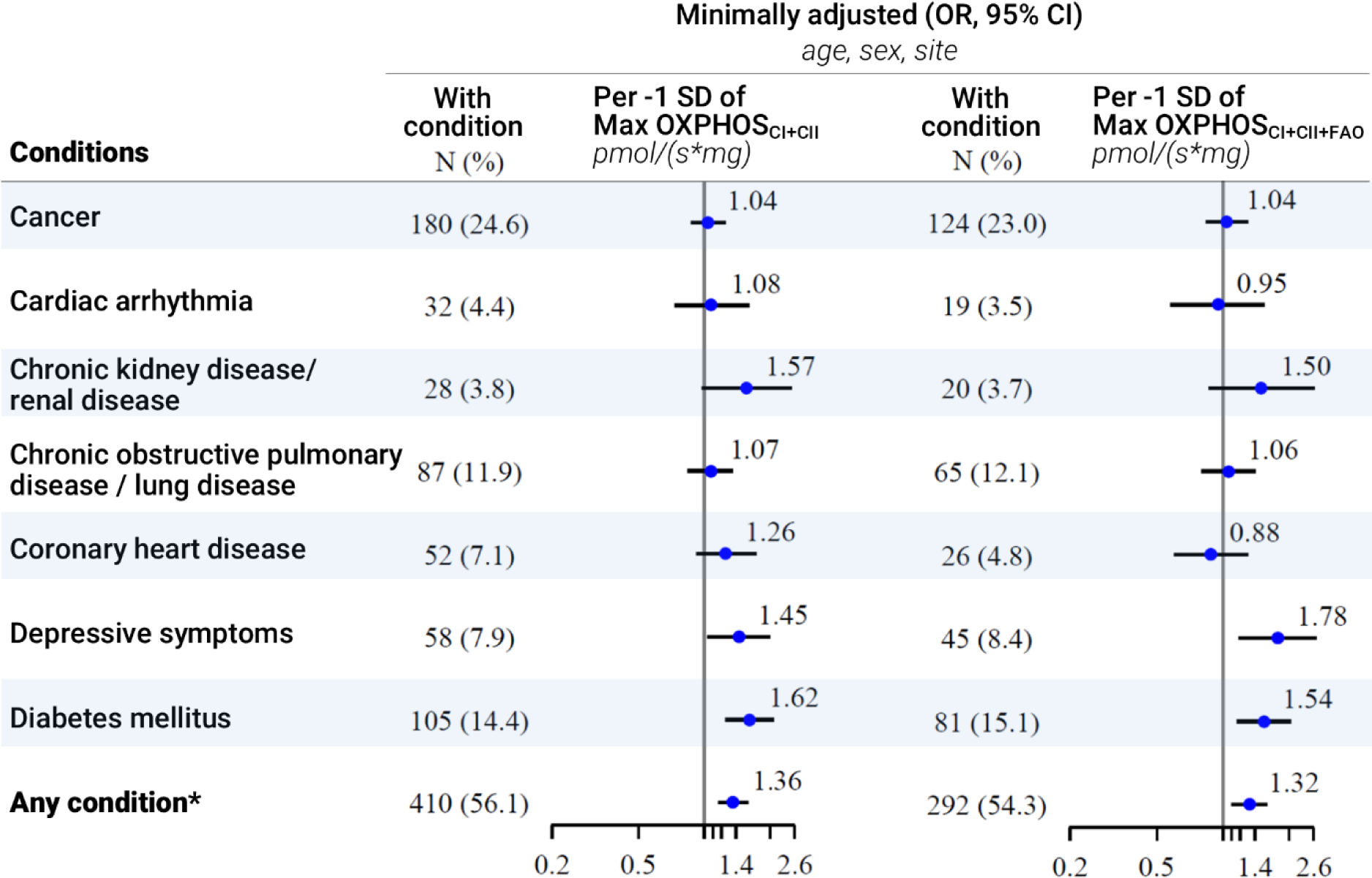
Associations of maximal muscle mitochondrial energetics and separate conditions from the multimorbidity index. Odds ratio (OR), the likelihood, of reporting a physician diagnosed condition for every 1 SD decrement of maximal muscle mitochondrial energetics is reported. *The OR for the likelihood of having 1 or more of the 11 conditions. Frequency of each condition are shown (N, %). Abbreviations: CKD: chronic kidney disease; COPD: chronic obstructive pulmonary disease; FAO: fatty-acid oxidation; OXPHOS: oxidative phosphorylation. Refer to **Supplementary Table 1** for variable names.

## Discussion

### Deficits in skeletal muscle mitochondrial bioenergetic capacities were associated with worse multimorbidity index scores

Declines in skeletal muscle mitochondrial energetics are linked to aging disease and dysfunction.^6,13–15,33^ Here, we characterized *ex vivo* skeletal muscle mitochondrial energetics assessed by high-resolution respirometry of permeabilized muscle fibers in a large group of clinically well-phenotyped older adults.^32^ Older adults with lower muscle mitochondrial energetics (supported by fatty and/or carbohydrates fuels) had greater multimorbidity index scores, and this was independent of age, sex, race, technician, education, smoking, alcohol, adiposity, and objectively-measured moderate-to-vigorous physical activity (MVPA). We also found that those with lower maximal muscle mitochondrial energetics (i.e., Max OXPHOS_CI+CII_ and Max OXPHOS_CI+CII+FAO_) were more likely to have some conditions of the index including diabetes mellitus, depressive symptoms, and chronic kidney disease, and perhaps other conditions where the associations did not individually reach statistical significance (i.e., cancer, cardiac arrhythmia, chronic obstructive pulmonary disease or lung disease, coronary heart disease) but had similar patterns (1.00<POR<1.40).

### Moderate-to-vigorous physical activity and adiposity do not explain the association between lower muscle mitochondrial energetics and greater multimorbidity burden

Older adults typically engage in less exercise as they age, possibly due to the higher burden of chronic health conditions. The overlapping effects of aging, adiposity, and exercise on skeletal muscle mitochondrial energetics are well-established.^25,31,34^ In studies of healthy older adults with no chronic conditions, reduction in physical activity, not chronological aging, negatively impacted muscle mitochondrial bioenergetics and oxidative capacity.^31,34,35^ One of these studies further demonstrated that decreased engagement in MVPA, not total activity (i.e., step counts per day), was associated with lower maximal muscle mitochondrial oxidative capacity.^31^ Increased adiposity has also been associated with reduced maximal skeletal muscle oxidative capacity.^30^ In our multivariable analyses, we adjusted for MVPA and adiposity, and lower muscle mitochondrial energetics remained significantly associated with greater multimorbidity burden. These results support that while MVPA and adiposity are known to influence muscle energetics, they do not entirely explain the associations between lower muscle mitochondrial energetics with greater multimorbidity burden. However, these outcomes do not discount the potential for exercise or physical activity interventions to improve muscle mitochondrial energetics^36–38^ and reduce the risk for multimorbidity. An important future question may be how various multimorbidity combinations influence physical activity and body composition in older adults.

### Deconstructing multimorbidity—energetically demanding diseases and tissues

Our multimorbidity index included chronic medical conditions primarily afflicting energetically demanding tissues—brain, skeletal muscle, heart, and kidney.^19^ Of the eleven deconstructed conditions, we were able to assess the associations of muscle mitochondrial energetics with seven conditions separately (cancer, cardiac arrhythmia, chronic kidney disease (CKD) or renal failure, chronic obstructive pulmonary or lung disease (COPD), coronary heart disease, depressive symptoms, and diabetes mellitus). SOMMA participants with lower maximal muscle mitochondrial energetics (Max OXPHOS_CI+CII_) had a higher likelihood of having diabetes mellitus (POR=1.62), chronic kidney disease (POR=1.57), or depressive symptoms (POR=1.45). This is consistent with other studies that have shown lower muscle mitochondrial energetics was associated with diabetes and chronic kidney disease.^24–27^ Some reports have linked depression, among other mood disorders, to mitochondrial dysfunction.^39–42^ This data is the first to show that those with lower muscle mitochondrial energetics had a greater likelihood of reporting depressive symptoms (CESD-10 ≥10) which aligns with a similar finding reported in peripheral blood mononuclear cells.^42^ Important to note, the outcomes between muscle mitochondrial bioenergetic capacities and depressive symptoms were minimally adjusted for only age, sex, and site. To better model the relationship, futures studies should assess potential confounders and mediators for both depressive symptoms and muscle mitochondrial energetics.

The multimorbidity index contains eleven age-related chronic conditions all with unique pathophysiology which may be differentially influenced by mitochondrial bioenergetic capacities. In fact, an in-depth review of the associations of skeletal muscle energetics with each condition may be difficult for one report to address. Further addressed in the discussion of limitations, but crucial to note here, is the enrollment selection bias that influenced the outcomes and sample sizes of chronic conditions we assessed. For example, the insignificant associations of muscle mitochondrial bioenergetics with cancer could be that muscle energetics differentially impact varied types of cancers.^43,44^ The types of cancers included in the index represent treated, surgically removed, or less aggressive forms of cancer (i.e., survivor bias) due to SOMMA enrollment criteria that excluded those with an active malignancy. We also observed patterns of associations between low muscle mitochondrial bioenergetics and coronary heart disease (POR=1.33) but not cardiac arrhythmia (POR=1.03). However, the associations with cardiac arrhythmia and coronary heart disease could be affected by selection bias in that we excluded individuals on chronic anticoagulation, which was mostly related to atrial fibrillation as well as anyone with a cardiovascular event in the prior 6 months. Both conditions contribute to heart failure which has been previously linked to reduced muscle mitochondrial oxidative capacity.^20–22^ Our data suggests no relationship between low muscle mitochondrial energetics and chronic obstructive pulmonary or other lung disease, which is similar to one study but incongruent with another.^45,46^ The composite multimorbidity index scores revealed that older adults with lower muscle mitochondrial energetics have a higher likelihood of greater multimorbidity burden.

When the index was deconstructed, the relationship between muscle mitochondrial energetics and each condition were of similar magnitude but not all statistically significant. Future studies of mitochondrial bioenergetics in both muscle and other tissues may reveal tissue-specific associations for various conditions.

### Limitations

The chronic conditions considered were based on self-report of a physician diagnosis with no information about disease duration nor severity. Nonetheless, prior research indicates that individuals could be relatively accurate in self-reporting physician diagnosed chronic conditions.^47,48^ Muscle mitochondrial energetics may be more relevant to diseases of the musculoskeletal system, whereas many conditions in our multimorbidity index are diseases that involve circulatory, respiratory, or nervous systems. For many of the chronic conditions included in the SOMMA multimorbidity index, we acknowledge that the influence is likely bidirectional. Chronic energetic demands and compounded damage from disease can result in loss of muscle mitochondrial bioenergetics such as reported in studies of cancer and diabetes.^24,49,50^ The impact of disease chronicity was also unaccounted for; a study using a rodent model of chronic kidney disease showed that uremic toxin accumulation was associated with the degree of decreased skeletal muscle oxidative capacity.^27^ Many prior studies have reported that those with a chronic aging-related condition tend to have lower muscle mitochondrial respiration, enzymatic activity, or capacities than those without the condition.^20–22,24–27^ Studies of blood cell bioenergetics which measure mitochondrial respiration in peripheral blood mononuclear cells and platelets may also reflect a more systemic measure of energetics than muscle.^51,52^ While SOMMA aimed to recruit older adults with a wide range of physical function, the exclusion of those with advanced chronic disease may pose a selection bias where only participants with combinations of multimorbidities who remain functional were enrolled, which is reflected in the relatively low prevalence of some conditions in SOMMA. This low prevalence prevented a complete assessment of some less common conditions. Importantly, this bias could also translate to an attenuation of the effect estimate: the reported associations here could be a conservative estimate of the true associations in the population. Finally, the study enrolled predominantly non-Hispanic White older adults which limits our ability to generalize to other diverse groups.

### Conclusions

Supporting the geroscience hypothesis, we showed that a biological pathway whose function is known to decline with age – poor muscle mitochondrial energetics – was associated with greater multimorbidity. Our data emphasize the relevance of muscle mitochondrial energetics to aging health and further support aging mitochondria as a therapeutic target.^33,53^ The impact of multimorbidity on healthcare systems and older adults is apparent in the tremendous costs of different disease combinations^54,55^ and the loss of health-span in aging populations.^56^ Developing a singular therapy targeting mitochondrial function that could improve multiple diseases at once would require more in-depth investigations of how muscle mitochondrial bioenergetic capacities are related to different chronic conditions. The future challenge in testing whether lower bioenergetics precede diagnosis of chronic diseases, besides longitudinal data, is that the severity and interactions of diseases may remain intertwined. These results suggest that a treatment to improve muscle mitochondrial energetics might have pluripotent effects on numerous disease outcomes.

## Acknowledgments

We acknowledge all SOMMA administrative, laboratory, clinical staff that have been listed previously.^32^

## Author Contributions

Using Contributor Roles Taxonomy (CRediT), TM led writing team for the original draft; TLB led formal analyses and managed data curation. Key conceptualization to this study involved PMCa, SRC, AJM, and GD. PMCa, PMCo, DEF, PAK, and ABN provided the most critical edits of the manuscript. SBK, SVR, BHG, FGST, KAD, LMS, and RTH reviewed and provided helpful comments to the manuscript. Finally, SRC, PMCa, ABN, and SBK enabled the study with either funding acquisition, project administration, supervision, and/or conceptualization to the study. All authors reviewed and approved of this manuscript.

## Data availability

This dataset is available upon formally requesting, accepting clinical data use agreements, and receiving approval at https://sommaonline.ucsf.edu/.

