## Supplementary Material for "Muscle mitochondrial bioenergetic capacities is associated with multimorbidity burden in older adults: the Study of Muscle, Mobility and Aging (SOMMA)"

**Supplementary Figure 1.** Flow diagram of analytic cohort

Enrolled in SOMMA.
(N=879)

Missing conditions: (N=5)

Refused CES-D: (N=9)

Completed medical history and CES-D. (N=865)

No respirometry assessed by protocols 1 nor 2: (N=101)

High-resolution respirometry performed.

(N=764)

Protocol 2 (N=540)

Protocol 1 (N=733)

| **Supplementary Table 1.**  Summary of high-resolution respirometry variables | |
| --- | --- |
| Abbreviation | Substrates |
| **Protocol 1** |  |
| Leak_CI_ | Pyruvate, malate |
| Submax OXPHOS_CI_ | ADP |
| OXPHOS_CI_ | ADP |
| Max OXPHOS_CI_ | Glutamate |
| Max OXPHOS_CI+CII_ | Succinate |
| Max ETS_CI+CII_ | FCCP* |
| **Protocol 2** |  |
| Leak_CI+FAO_ | Palmitoylcarnitine, malate |
| OXPHOS_CI+FAO_ | ADP |
| Max OXPHOS_CI+FAO_ | Glutamate |
| Max OXPHOS_CI+CII+FAO_ | Succinate |

*Notes.* Summary of high-resolution respirometry variables, complete methods found in online methods. The abbreviations of respirometry measure appears on the left column; the substrates added sequentially prior to the measure is on the right column.

*Abbreviations:* CI, complex I-supported; CII, complex II-supported; FAO, fatty acid oxidation; OXPHOS, oxidative phosphorylation; ETS, electron transport system; ADP, adenosine diphosphate; submax, submaximal; max, maximal. *Carbonyl cyanide-p-trifluoromethoxyphenylhydrazone (FCCP) elicits uncoupled maximal respiratory capacity of ETS.
